# Intimate partner violence victimization and perpetration with greater probability of brain injury in victims: A preliminary study

**DOI:** 10.64898/2026.11.15.26368160

**Authors:** Shambhu P. Adhikari, Caroline McLennan, Krystal Rothlander, Karen Mason, Paul van Donkelaar

## Abstract

**Background:** Intimate Partner Violence (IPV) can be either unidirectional or bidirectional. At times when it is bidirectional, the impact of the IPV would likely be greater in women compared to men especially in cases involving physical IPV. This study compared IPV victimization and perpetration characteristics and explored the relationship between IPV victimization and brain injury (BI) among women victims of IPV.

**Methods:** In this cross-sectional study, 23 women (IPV victims) aged 18-60 years who experienced at least one reported incident of IPV-BI were recruited from different community-based, women-serving organizations. Data collection included a comprehensive assessment of IPV, BI, and psychopathological comorbidities. The Conflict Tactics Scale-2 (CTS2) was used to assess for victimization and perpetration.

**Results:** The participants had significantly lower scores on the physical assaults (t = 5.94, p < 0.0001, η_p_^2^ = 0.62), and higher scores on the injury (t = 6.12, p < 0.001, η_p_^2^ = 0.63) domains of the CTS2 compared to the perpetrator/partner. The physical assault scores of the perpetrator/partner were significantly correlated with the injury scores of the victim (r = 0.86, p < 0.001). Moreover, victims’ injury scores and their BISA scores were also significantly correlated (r = 0.69, p = 0.002).

**Conclusions:** Women victims of IPV, in contrast to perpetrator/partners, demonstrated greater physical injury, which were associated with an increased likelihood of BI in this cohort. Findings should be interpreted cautiously, as partners’ data were based on victim reports. Further studies are warranted to explore the possible occurrence of BI in perpetrators and then directly compare the injury exposure in victims versus perpetrators. Based on the findings of this study, it is clear there is a higher prevalence, as well as greater exposure, of BI and more serious consequences of IPV and/or IPV-BI on victims compared to that of perpetrators.

## Introduction

Intimate partner violence (IPV) is a recognized global public health problem (1) that affects more than 4 in 10 women in Canada (2). IPV is violence or harm caused by a current or former intimate partner. Survivor of IPV (“victim” is used hereafter) refers to a person who experienced non-fatal IPV. Perpetrator of IPV (the term “perpetrator” is used hereafter) refers to a person who abused an intimate partner (3). It is well established that women report a high prevalence of IPV, though individuals of all genders experience it (4).

Due to the unique nature of IPV and challenges for collection of data directly from perpetrators, studies that assess both IPV victimization and IPV perpetration in the same sample are sparse. The likelihood of perpetrating IPV without being a victim at the same time is higher in men compared to women (5). Men usually tend to under-report perpetration whereas women tend to under-report victimization (5). Therefore, the prevalence of IPV victimization (approximately 27.0 – 37.2%) and IPV perpetration (approximately 12.9 to 24.8%) reported elsewhere (6, 7) may be underestimated.

One of the most understudied consequences of IPV is brain injury (BI) from physical abuse. In a scoping review, Campbell et. al., (3) reported that BI is prevalent in both victims and perpetrators of IPV. Furthermore, there is a growing body of literature reporting an association between physical IPV and BI (3, 8–12). However, there is scarce evidence linking the level of physical assault by perpetrators with the injury level experienced by victims in the same sample. Moreover, there is a lack of literature that directly examined the association between IPV victimization and BI due to IPV (IPV-BI). Therefore, this study had two primary objectives: 1) to compare the IPV characteristics (i.e. victimization and perpetration) among victims and perpetrators of IPV in the same sample, and 2) to directly assess BI in victims using an IPV-BI screening tool, in order to explore the potential linkage between the IPV victimization and IPV-BI occurrence in IPV victims. We hypothesized IPV victims would experience higher levels of physical injury, and thereby would increase the likelihood of acquiring BI compared to perpetrators.

Given the established relationship between IPV exposure and adverse mental health outcomes, a post-hoc secondary analysis was undertaken to examine the associations of post-traumatic stress disorder (PTSD), depression, and anxiety with the various domains of the conflict tactic scale (CTS2) while controlling for potential covariates, including experience of non-fatal strangulation, number of non-IPV-related brain injuries, age, ethnicity, and income. Exploring the associations between PTSD, depression, anxiety symptoms, and CTS2 domain was considered important to better understand the psychological correlates of IPV while accounting for relevant demographic and injury-related factors.

## Materials and Methods

Ethical approval to conduct this study was received from the clinical research ethics board at the University of British Columbia (UBC CREB numbers H16-02792, H20-02412). All participants provided written, informed consent prior to participation. Research coordinators described all aspects of the study to the participants prior to obtaining written, informed consent. Data collection was conducted from February 2021 through November 2025.

### Design

A cross-sectional study.

### Participants and recruitment

Women (IPV victims) aged 18-60 years who experienced at least one reported incident of IPV-BI was the principal criterion for the study. However, participants were not excluded if they had experienced BI outside the context of IPV in order to recruit participants with a spectrum of previous BIs. As such, 23 women were recruited from different community-based, women-serving organizations in and around Kelowna, British Columbia, Canada.

### Measures

As described in one of the published papers by the research team (13), data collection included a comprehensive assessment of IPV, BI, and psychopathological comorbidities. Various demographic and clinical characteristics of the victim such as age, ethnicity, annual income, previous history of BI from non-IPV causes were also collected.

#### Revised Conflict Tactic Scale (CTS2)

The CTS2 (14) is a commonly used, reliable and valid tool for measuring the prevalence of victimization and perpetration in IPV victims (15). This tool was designed to be an objective measure for exploring the prevalence of IPV during the previous 12 months. The respondent is asked to report on both their own use of a variety of tactics as well as whether their partner used those tactics (i.e., ‘I slammed my partner against a wall.’ ‘My partner slammed me against a wall’). The CTS2 consists of 78 items, with 39 items (all odd-numbered questions) assessing participants’ own behaviors and experiences, and 39 corresponding even-numbered items assessing those of their intimate partners. Consistent with the CTS2 administration guidelines, participants are explicitly instructed that some questions refer to themselves, whereas others refer to their partners. Accordingly, IPV victims completed all CTS2 items during a single assessment session, providing information on both their own experiences and their partners’ behaviors. This approach generates separate scores for victimization and perpetration across each CTS2 domain.

The CTS2 has been widely used in research, evidence-based initiatives to inform IPV treatment and policies, therapy to assist disclosure, and to monitor behavioral and treatment progress, (16, 17) and is the only measure capturing the extent of multiple forms of victimization and perpetration (18). It is a 39-item, self-report questionnaire with five subscales; 1) negotiation (6 items), 2) psychological aggression (8 items), 3) physical assault (12 items), 4) injury (6 items), and 5) sexual coercion (7 items). Respondents are asked to give a score on both victimization and perpetration using a Likert scale. Accordingly, participants who experienced IPV completed the CTS2 for both themselves and their partners, generating separate scores for victims and partners as recommended by the standardized administration guidelines of the instrument in the current study. The response options are: 1: once; 2: twice; 3: 3–5 times; 4: 6–10 times; 5: 11–20 times; 6: >20 times; 7: not in referent period but happened before; and 0: never (14). Studies have analyzed data using either the raw scores directly or using the mid-point scores (19, 20). Consistent with Straud and coworkers (19), we used raw scores for analysis in the current study.

#### Brain Injury Severity Assessment (BISA) interview

The BISA (21) was used to assess IPV-BI. It is a semi-structured interview containing a series of questions regarding loss of consciousness, dizziness, memory loss, feeling stunned or disoriented, or seeing stars or spots following potential traumas to the brain. The BISA provides a summary score ranging from 0 to 8. Any score between 1 and 8 indicates the occurrence of at least one probable BI. It consists of three sub-scales; 1) frequency - an estimate of the number of previous potential BIs (0-4 scale), 2) recency - the time since the most recent event resulting in a potential BI (0-3 scale), and 3) severity - indicates whether a loss of consciousness and/or post-traumatic amnesia ever occurred as a result of IPV (0-1 scale). Although it has not yet been characterized psychometrically, it has been widely used in IPV-BI research (9, 21, 22).

#### Assessment of Depression, Anxiety and Post-traumatic Stress Disorder (PTSD)

The Patient Health Questionnaire (PHQ-9) (23), Generalized Anxiety Disorder Scale (GAD-7) (24), and PTSD Checklist for DSM-5 (PCL-5) (25) were used to measure depression, anxiety and PTSD, respectively. Further details on each of the measures used in this study are provided in a previously published protocol paper by the research team (13).

### Statistical Analysis

Descriptive statistics [(mean ± SD or number (percentage)] were conducted to analyze the demographic and clinical characteristics of the cohort. The Kolmogorov-Smirnov (K-S) test revealed the outcome variables were normally distributed (p > 0.05), and, thus, parametric tests were used. Since, victims provided responses for themselves and for their partners, paired t-tests were conducted to compare CTS-2 victim and partner scores on various domains. Post-hoc secondary analysis was conducted to explore any association between the injury scores of the victim with their BISA scores and the physical assault scores of the partners. Additionally, multi-step multiple linear regression analysis was conducted to explore any relationship among different exposure variables (GAD-7, PHQ-9, PCL-5, experience of non-fatal strangulation, number of non-IPV related BIs, age, ethnicity, and income) with the five different domains of the CTS2 (negotiation, psychological aggression, physical assault, injury and sexual coercion). The partial Eta squared (η_p_^2^) was calculated to determine whether the effect sizes were small (0.01), medium (0.06), or large (0.14) (26). Significance was set at *p* < 0.05. All statistical analyses were performed using SPSS Statistics version 30.0 (IBM Corp, Armonk, NY), and GraphPad-Prism software, version 10.4.0 (Dotmatics, Boston, MA).

## Results

In total, 23 participants (all were women who had experienced IPV) completed the CTS2, psychopathological and demographics questionnaire. Among them, only 20 participants agreed to complete the BISA interview. Regarding demographic and clinical characteristics, five participants did not disclose information on non-IPV related BIs and one participant did not provide information on annual income. Detailed characteristics of the participants are depicted in **Table 1**. The victims of IPV completed the CTS2 questionnaire for both victim and partner and provided separate raw scores for victims as well as partners, which are outlined in **Table 1**.

**Table 1:**
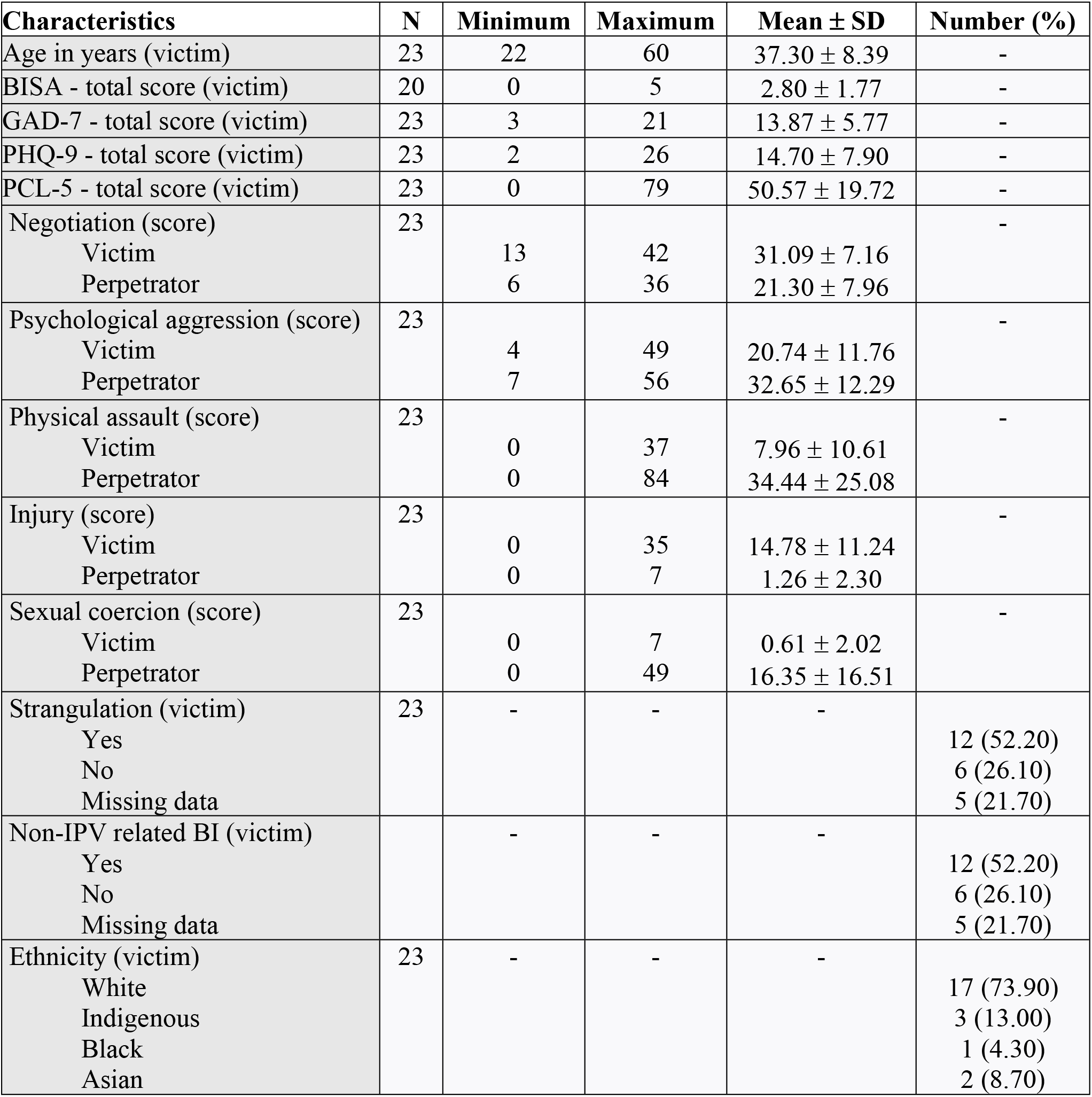
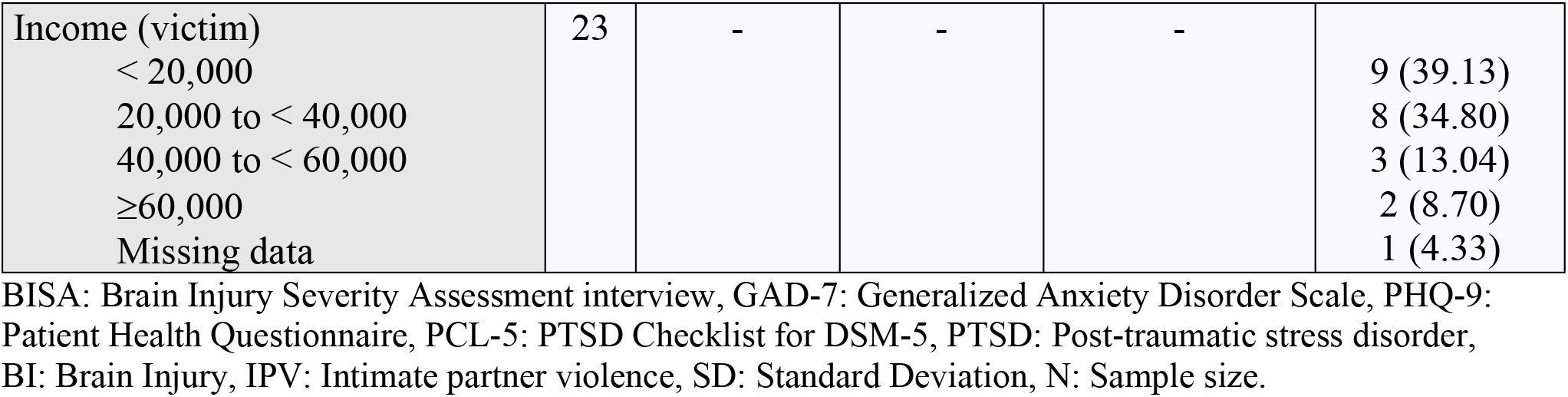
Characteristics of the participants.

| **Characteristics** | **N** | **Minimum** | **Maximum** | **Mean ± SD** | **Number (%)** |
|---|---|---|---|---|---|
| Age in years (victim) | 23 | 22 | 60 | 37.30 ± 8.39 | - |
| BISA - total score (victim) | 20 | 0 | 5 | 2.80 ± 1.77 | - |
| GAD-7 - total score (victim) | 23 | 3 | 21 | 13.87 ± 5.77 | - |
| PHQ-9 - total score (victim) | 23 | 2 | 26 | 14.70 ± 7.90 | - |
| PCL-5 - total score (victim) | 23 | 0 | 79 | 50.57 ± 19.72 | - |
| Negotiation (score) | 23 |  |  |  | - |
| Victim |  | 13 | 42 | 31.09 ± 7.16 |  |
| Perpetrator |  | 6 | 36 | 21.30 ± 7.96 |  |
| Psychological aggression (score) | 23 |  |  |  | - |
| Victim |  | 4 | 49 | 20.74 ± 11.76 |  |
| Perpetrator |  | 7 | 56 | 32.65 ± 12.29 |  |
| Physical assault (score) | 23 |  |  |  | - |
| Victim |  | 0 | 37 | 7.96 ± 10.61 |  |
| Perpetrator |  | 0 | 84 | 34.44 ± 25.08 |  |
| Injury (score) | 23 |  |  |  | - |
| Victim |  | 0 | 35 | 14.78 ± 11.24 |  |
| Perpetrator |  | 0 | 7 | 1.26 ± 2.30 |  |
| Sexual coercion (score) | 23 |  |  |  | - |
| Victim |  | 0 | 7 | 0.61 ± 2.02 |  |
| Perpetrator |  | 0 | 49 | 16.35 ± 16.51 |  |
| Strangulation (victim) | 23 | - | - | - |  |
| Yes |  |  |  |  | 12 (52.20) |
| No |  |  |  |  | 6 (26.10) |
| Missing data |  |  |  |  | 5 (21.70) |
| Non-IPV related BI (victim) |  | - | - | - |  |
| Yes |  |  |  |  | 12 (52.20) |
| No |  |  |  |  | 6 (26.10) |
| Missing data |  |  |  |  | 5 (21.70) |
| Ethnicity (victim) | 23 | - | - | - |  |
| White |  |  |  |  | 17 (73.90) |
| Indigenous |  |  |  |  | 3 (13.00) |
| Black |  |  |  |  | 1 (4.30) |
| Asian |  |  |  |  | 2 (8.70) |
| Income (victim) | 23 | - | - | - |  |
| < 20,000 |  |  |  |  | 9 (39.13) |
| 20,000 to < 40,000 |  |  |  |  | 8 (34.80) |
| 40,000 to < 60,000 |  |  |  |  | 3 (13.04) |
| ≥60,000 |  |  |  |  | 2 (8.70) |
| Missing data |  |  |  |  | 1 (4.33) |

BISA: Brain Injury Severity Assessment interview, GAD-7: Generalized Anxiety Disorder Scale, PHQ-9: Patient Health Questionnaire, PCL-5: PTSD Checklist for DSM-5, PTSD: Post-traumatic stress disorder, BI: Brain Injury, IPV: Intimate partner violence, SD: Standard Deviation, N: Sample size.

All victims endorsed one or more of the CTS2 domains by self and partner. The prevalence of victimization and perpetration reported by the victims, expressed as a percentage, is shown in **Table 2**. One of the interesting findings in **Table 2** is the percentage of participants endorsing one or more items in the injury domain of victims was equal to the percentage of participants who reported physical assault by their partners. Participants also reported the extent of sexual coercion by their partner was more than nine times higher than that undertaken by them (**Table 2**).

**Table 2:** Revised Conflict Tactic (CTS2) scale: Prevalence of various domains, N = 23.

| **CTS2 domains** | **Victim** |  | **Perpetrator** |  |
|---|---|---|---|---|
|  | **n** | **%** | **n** | **%** |
| Negotiation | 23 | 100% | 23 | 100% |
| Psychological aggression | 23 | 100% | 23 | 100% |
| Physical assault | 16 | 69.57% | 22 | 95.65% |
| Injury | 22 | 95.65% | 16 | 69.57% |
| Sexual coercion | 2 | 8.70% | 17 | 73.91% |

CTS2: Revised Conflict Tactic scale; n: number of participants in each domain; N: Total sample

Means, standard deviations, and score ranges for victims and partners on the CTS2 domains are presented in **Table 1**. Paired t-tests revealed that victims had significantly lower scores on the physical assaults (t = 5.94, df = 22, p < 0.0001, η_p_^2^ = 0.62), psychological aggression (t = 6.43, df = 22, p < 0.0001, η_p_^2^ = 0.65), and sexual coercion (t = 4.83, df = 22, p < 0.0001, η_p_^2^ = 0.52) domains (Figure 1: A, B, and C), and higher scores on the negotiation (t = 5.12, df = 22, p < 0.0001, η_p_^2^ = 0.55), and injury (t = 6.12, df = 22, p < 0.0001, η_p_^2^ = 0.63) domains (Figure 1: D, and E). A large effect size, ranging from 0.52 to 0.65 was found in these comparisons.

**Figure 1:**
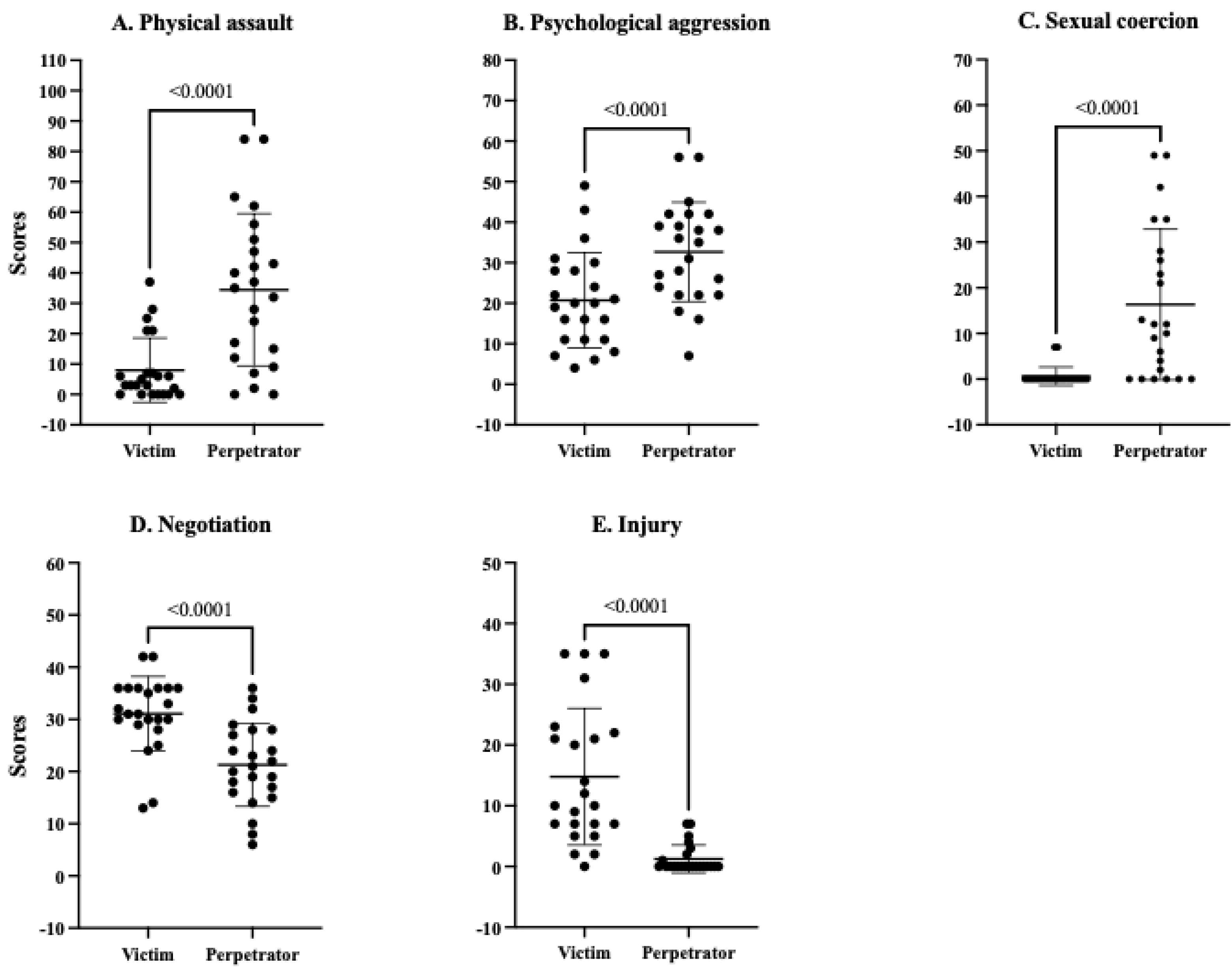
Comparison between victim and perpetrator on the various domains of the Conflict Tactic Scale-2 (CTS-2). A) Physical domain, B) Psychological aggression domain, C) Sexual coercion domain, D) Negotiation domain, and E) Injury domain.

The physical assault scores of the partner were significantly correlated with the injury scores of the victim (R^2^ = 0.73, r = 0.86, p < 0.001) (**Figure 2, A**). The higher the level of physical assault from the partner, the higher was the injury level in the victim. Consistently, the injury scores of the victim were significantly correlated with their BISA scores (R^2^ = 0.48, r = 0.69, p = 0.002) (**Figure 2, B**). The higher the injury level in the victim, the higher their BISA scores.

**Figure 2:**
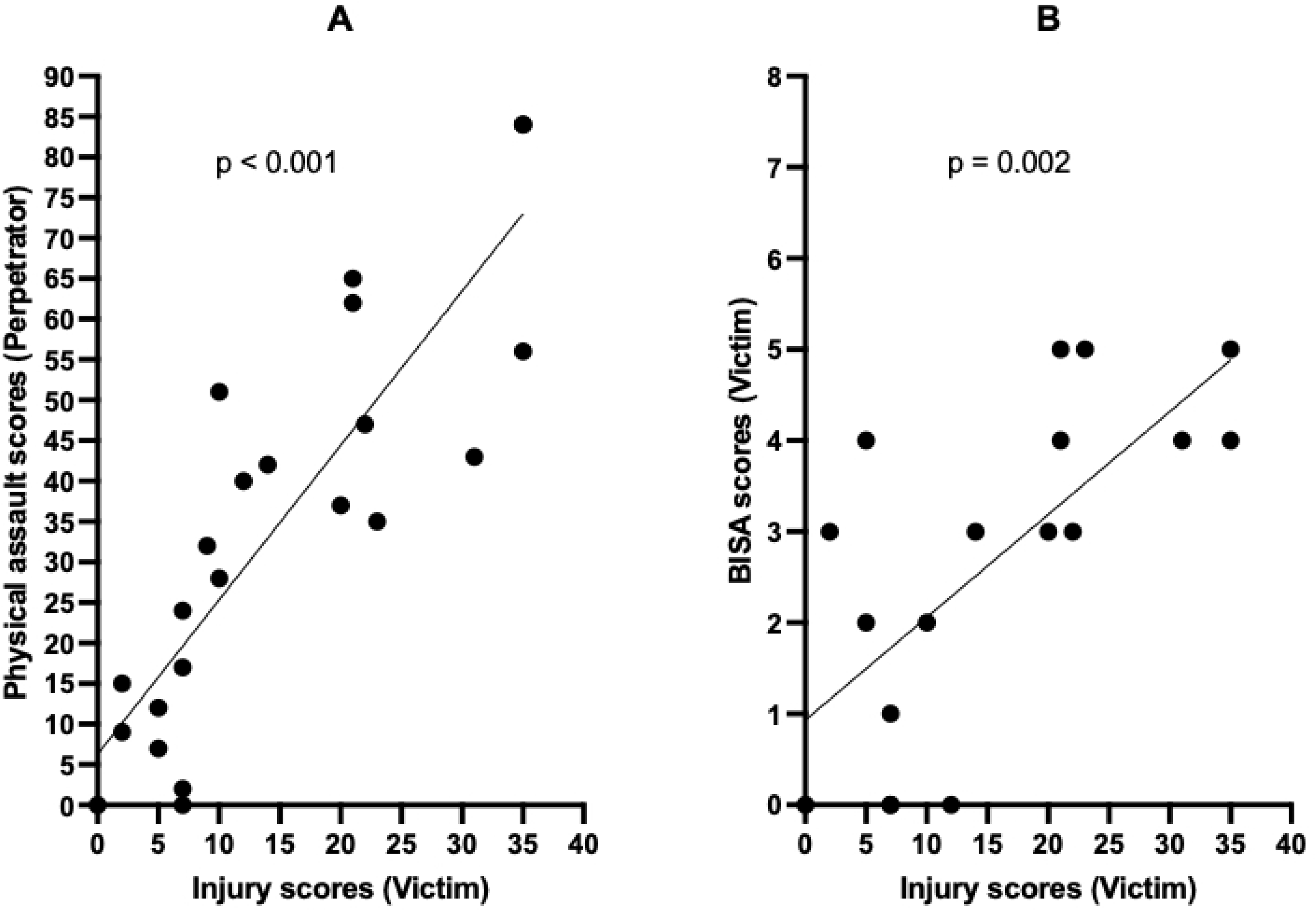
Outcome of the correlation analysis. A) Correlation between the physical assault scores of the perpetrator versus injury scores of the victim, B) Correlation between the injury scores of the victim versus BISA scores of the victim. BISA: Brain injury severity assessment tool.

The multi-step multiple linear regression analysis undertaken to address the post-hoc secondary objective revealed that depression (measured using the PHQ-9) and PTSD (measured using the PCL-5) were significantly associated with the negotiation domain of the CTS2 (**Table 3, model 1**). This association still persisted even after accounting for other covariates (experience of non-fatal strangulation, number of non-IPV related BI, age, ethnicity, and income) (**Table 3, models 2 and 3**). The results showed that the negotiation domain was negatively impacted by the presence of PTSD symptoms (Coefficient: -0.37, R^2^: 0.74, t = -3.01, p = 0.02, 95% CI: -0.66, -0.09), and positively impacted by the presence of depression symptoms (Coefficient: 0.99, R^2^: 0.74, t = 3.51, p < 0.01, 95% CI: 0.34, 1.65). We did not find any association (p > 0.05) of any other exposure variables (GAD-7, PHQ-9, PCL-5, experience of strangulation, number of non-IPV related BI, age, ethnicity, and income) with any domains of the CTS2 (psychological aggression, physical assault, injury and sexual coercion) (**Table 3**).

**Table 3:** Multi-steps multiple linear regression showing the relationships among various exposure variables and the CTS2 domains.

| Models |  | Negotiation |  | Psychological aggression |  | Physical assault |  | Injury |  | Sexual coercion |  |
|---|---|---|---|---|---|---|---|---|---|---|---|
|  |  | Coefficients (B) | Sig. | Coefficients (B) | Sig. | Coefficients (B) | Sig. | Coefficients (B) | Sig. | Coefficients (B) | Sig. |
| 1 | GAD-7 - total score | 0.09 | 0.82 | 0.49 | 0.57 | - 0.19 | 0.75 | 0.85 | 0.37 | 0.04 | 0.79 |
|  | PHQ-9 - total score | 0.95 | **< 0.01**\* | 0.99 | 0.08 | 0.56 | 0.15 | 0.32 | 0.58 | 0.05 | 0.56 |
|  | PCL-5 - total score | - 0.37 | **0.01**\* | - 0.50 | 0.09 | - 0.23 | 0.25 | - 0.33 | 0.28 | 0.01 | 0.99 |
| 2 | GAD-7 - total score | 0.25 | 0.53 | 0.83 | 0.36 | 0.09 | 0.87 | 1.03 | 0.31 | 0.07 | 0.65 |
|  | PHQ-9 - total score | 0.86 | **< 0.01**\* | 0.83 | 0.15 | 0.42 | 0.26 | 0.21 | 0.73 | 0.03 | 0.72 |
|  | PCL-5 - total score | - 0.37 | **< 0.01**\* | - 0.51 | 0.08 | - 0.24 | 0.21 | - 0.33 | 0.29 | 0.00 | 0.99 |
|  | Strangulation (Yes/No) | 2.74 | 0.30 | 1.65 | 0.78 | 2.07 | 0.60 | 6.88 | 0.31 | 0.42 | 0.68 |
|  | # of non-IPV related BI | 0.76 | 0.26 | 1.90 | 0.21 | 1.56 | 0.13 | 0.66 | 0.69 | 0.17 | 0.49 |
| 3 | GAD-7 - total score | 0.18 | 0.69 | 0.34 | 0.76 | 0.29 | 0.70 | 1.35 | 0.24 | 0.18 | 0.30 |
|  | PHQ-9 - total score | 0.99 | **< 0.01**\* | 0.96 | 0.18 | 0.30 | 0.52 | - 0.28 | 0.68 | 0.00 | 0.99 |
|  | PCL-5 - total score | - 0.37 | **0.02**\* | - 0.54 | 0.09 | - 0.21 | 0.31 | - 0.29 | 0.35 | 0.01 | 0.85 |
|  | Strangulation (Y/N) | 1.42 | 0.64 | 4.41 | 0.53 | 1.29 | 0.79 | 9.92 | 0.18 | - 0.29 | 0.79 |
|  | # of non-IPV related BI | 1.47 | 0.18 | 0.08 | 0.97 | 2.33 | 0.19 | - 0.45 | 0.85 | 0.68 | 0.09 |
|  | Age (yrs) | - 0.11 | 0.58 | - 0.45 | 0.33 | 0.01 | 0.96 | 0.10 | 0.82 | 0.05 | 0.46 |
|  | Ethnicity (categorical) | - 0.07 | 0.97 | - 0.11 | 0.98 | -3.03 | 0.36 | -5.76 | 0.25 | - 0.77 | 0.29 |
|  | Income (categorical) | -1.80 | 0.41 | 4.62 | 0.36 | 0.90 | 0.79 | 8.39 | 0.13 | - 0.56 | 0.47 |

GAD-7: Generalized Anxiety Disorder Scale, PHQ-9: Patient Health Questionnaire, PCL-5: PTSD Checklist for DSM-5, PTSD: Post-traumatic stress disorder, BI: Brain Injury, IPV: Intimate partner violence; \* significant at p< 0.05.

## Discussion

Given the wide use of the CTS2 in estimating the prevalence of IPV victimization and perpetration, this study was designed to use the CTS2 to explore an association between IPV and IPV-BI by correlating it with scores from the BISA tool. The major findings of the study were: 1) all participants endorsed violence by self and partner, however they demonstrated a higher level of victimization on CTS2 compared to perpetration, 2) the higher the level of physical assault by perpetrators, the higher the injury level on the victims, and 3) the level of injury in victims measured by CTS2 was directly associated with potential BI exposure measured using the BISA, indicating the occurrence of BI on victims.

Given the unique nature of IPV, studies that have enrolled both members of known victim-perpetrator couples are rare (27) due to ethical, safety, and confidentiality concerns. One notable exception is Gondolf’s multi-site evaluation of court-referred batterer intervention programs, which collected follow-up data from both identified perpetrators and their female partners (28). However, this study was primarily designed to evaluate intervention outcomes rather than to conduct dyadic analyses. In contrast, dyadic IPV studies such as those conducted by Capinha et al. (29), and Lefebvre et al. (30) recruited both partners and assessed perpetration and victimization from each partner’s perspective; however, participants were not necessarily enrolled as pre-identified victim-perpetrator couples. Consequently, research involving both members of known victim-perpetrator couples remains limited, and studies comparing physical injury outcomes, including probable brain injury, between victims and perpetrators within the same relationship remain scarce. In the current study, victim participants provided information regarding both their own experiences and their partners’ behaviors. Therefore, the findings of the present study should be interpreted with caution.

IPV can be either unidirectional and bidirectional (31–34). Findings of the current study demonstrated bidirectional IPV because all women in this study confirmed violence by self and partner. The impact of IPV on women was more serious than on men, however, because women victims in this study in contrast to men perpetrators demonstrated probably BI due to the violence. Importantly, women’s use of violence may not reflect aggression or intent to harm, but rather self-defence. Many women may respond physically to protect themselves from assaults perpetrated by their partners. This defensive violence is often aimed at stopping or escaping an attack and typically results in less severe physical injury than the violence inflicted by male perpetrators. In contrast, violence used by perpetrators is more likely to be offensive and cause serious harm. Even so, bidirectional IPV is especially concerning because when women defend themselves with violence, it can increase the risk of further retaliation and repeated victimization (35). This creates a cycle in which women are exposed to ongoing violence, raising the likelihood of repeated BIs and more serious short- and long-term health consequences.

On the other hand, the exposure to violence experienced by perpetrators would have been underestimated in the current study because perpetrators were not directly interviewed. Instead, the perpetrators’ scores on the CTS2 were provided by the victims. The prevalence of BI (either TBI or non-fatal strangulation-related alteration in consciousness) in both victims (average: 66%, range from 43% to 89%) and perpetrators (average: 40%, range from 5% to 75%) is usually higher than in that in the general population (18%) (3, 36). Further studies are required to explore the occurrence of BI in perpetrators with methodological rigor and directly compare the exposure to violence and severity of injury in victims and perpetrators. However, based on the findings of this study, we can argue that BI as a result of IPV would be much more likely in women victims compared to men perpetrators.

Additionally, we also explored the relationships between various psychopathological measures and different components of the CTS2 during the post-hoc secondary analysis. We found that psychological distress, mainly PTSD and depression, modulated the negotiation component of the CTS2. In particular, the higher the level of PTSD symptoms, the lower the scores on the negotiation domain. This finding makes sense because mental health issues often lead to difficulty in the cognitive decision-making process (37). By contrast, the higher the scores on the negotiation domain, the higher the levels of depression. This could be because people with depression may use negotiation strategies more frequently to prevent escalation, attempt to smooth over disagreements to avoid stress, or overestimate the amount of effort they put into resolving conflict. Thus, the reasons behind the positive association between the negotiation and depression demonstrated in the current study is not clear enough and warrants further research. More generally, low levels of negotiation among the victims and perpetrators might have aggravated the level of violence in this cohort. This might have led to higher levels of physical assault from the perpetrators, subsequently resulting in higher prevalence of BI in women victims. However, the directionality of this relationship is not clear, and more robust studies are warranted to explore the intersection between mental health comorbidities and IPV-BI.

### Limitations

There are a number of limitations to this study. First, as mentioned elsewhere in this paper, perpetrators were not directly interviewed. Victims provided responses for themselves and for their partners. Therefore, the perpetrators’ response might have been underscored whereas the victims’ response might have been overscored. It would be ideal if the prevalence of IPV in victims and perpetrators could be explored by directly interviewing both parties. For example, Babcock and colleagues (38) used phone screening to examine past violence or dissatisfaction amongst couples with a history of conflict. Although the study excluded couples who felt unsafe participating, underreporting may still be present due to fear of retaliation, partner’s presence during the call, and shame or stigma. Additionally, couples experiencing more severe, coercive, or controlling violence may be less likely to respond, meaning the sample might have overrepresented those at lower to moderate risk or those more comfortable disclosing (see also e.g., Capinha et al., 2024; Marshall et al., 2025). To summarize, couple-based IPV studies often end up overrepresenting lower-risk relationships because partners experiencing severe or coercive violence are less likely to participate, especially when recruitment involves joint screening or disclosure of past aggression. In reality, however, due to the unique nature of the violence, victims’ safety issues, etc., it is challenging to interview both victims and perpetrators, though allowing each partner to enroll independently rather than as a pair might help minimize some biases. Some robust methodology might be required for exploring the true prevalence of victimization and perpetration, particularly in perpetrators.

Second, the study identified a significant association between the victims’ injury scores and their exposure to probable BI. Perpetrators’ BI could not be assessed because they were not enrolled in this study. However, based on the findings in a review by Campbell et al., (3) perpetrators might have acquired some level of BI. Therefore, further studies are warranted to explore the possible occurrence of BI in perpetrators and then directly compare the exposure of injury in victims versus perpetrators. Based on the available evidence (3, 35) and the findings of the current study, we would argue for higher prevalence of BI, greater exposure of BI and more serious consequences of IPV and/or IPV-BI on victims compared to that on perpetrators.

Third, the sample size in this study was relatively small. This paper was part of a larger study and included only preliminary findings. A larger sample, including a more heterogenous population in terms of gender, education, ethnicity, geographical regions, socioeconomical status, etc. would probably provide additional information.

## Conclusion

Women who experienced IPV reported significantly higher levels of physical injury than male perpetrators, with greater injury severity being associated with BI in this cohort. Given that partner behaviors were reported by victims rather than perpetrators themselves, the study findings should be interpreted with caution. More research is needed to accurately estimate the prevalence of BI on victims as well as on perpetrators and then to directly compare the load of injury among victims and perpetrators in IPV.

## Acknowledgements

We wish to acknowledge the community organizations who supported this project through promotion, recruitment, and execution, including Central Okanagan Elizabeth Fry Society (Kelowna, B.C.), Connect Counselling and Therapy (Kelowna, B.C.), William and Associates Counselling Services (Kelowna, B.C.), Kelowna Women’s Shelter (Kelowna, B.C.), and Mamas for Mamas (Kelowna, B.C.). We extend special gratitude to the valued, lived experience survivors of IPV-BI who supported this project with their time, energy and attention as research participants. Finally, we would like to show our deepest gratitude to all the individuals and the organizations who directly or indirectly worked with us to accomplish this study.

## Authors’ Contributions

**SPA**: Conceptualization, methodology, data analysis, writing (original draft), writing (review and editing).

**CMcL, and KK**: Data collection, writing (review and editing).

**KM:** Conceptualization, methodology, writing (review and editing).

**PVD:** Conceptualization, methodology, supervision, writing (review and editing).

## Ethical Considerations

The University of British Columbia clinical research ethics board granted approval for this research study (UBC CREB numbers H16-02792, H20-02412).

## Consent to Participate

All participants gave written, informed consent to participate in the study before taking part.

## Consent for Publication

All participants and authors of this study provided consent for publication. Non-essential identifying details of the participants have been omitted.

## Availability of Data and Material

Anonymized data is available upon request from the corresponding author.

